# B-Mode ultrasound patterns and findings of the liver among hepatitis B and C patients: a cross-sectional study in a Rwandan tertiary hospital

**DOI:** 10.1101/2025.11.05.25339480

**Authors:** Florien Ujemurwego, Rachel Ankunda, Gaspard Harerimana, Placide Bikorimana, Josiane Uwineza, Jean de Dieu Niyonteze

## Abstract

**Background:** Hepatitis B virus (HBV) and hepatitis C virus (HCV) infections remain major global health challenges, with Rwanda reporting prevalence rates of 2.1% and 6.7%, respectively. Ultrasonography is crucial in assessing chronic viral hepatitis; however, data on sonographic liver patterns among HBV and HCV patients in Rwanda are limited.

**Objectives:** To identify B-mode ultrasound liver patterns among hepatitis B and C patients and assess their association with sociodemographic factors.

**Method:** A cross-sectional study was conducted among 240 patients with viral hepatitis (120 HBV, 120 HCV). B-mode ultrasound findings were analyzed for associations with demographic variables.

**Results:** Among 240 participants (mean age 52.7 ± 14.9 years; 57.1% male), the most frequent liver findings were patterns of chronic liver alteration suggestive of fibrosis (21.7%), fatty liver (8.8%), suspicious nodules (5.0%) and hepatomegaly (2.9%). Patterns of chronic liver alteration were significantly more common in HCV than HBV (14.2% vs. 7.5%, p = 0.012) and were associated with older age (p < 0.001) and female gender (p = 0.006).

**Conclusion:** Ultrasound revealed distinct liver morphological changes among hepatitis B and C patients, more frequent in HCV, older participants, and females, suggesting differences in infection duration or healthcare-seeking behavior. Ultrasound remains a practical tool for identifying and monitoring liver changes in resource-limited settings.

**Contribution:** This study provides real-world data on sonographic liver patterns in hepatitis B and C, reinforcing ultrasound’s value as an accessible diagnostic tool in African radiology practice

## 1. Introduction

Hepatitis B virus and hepatitis C virus infections are major global public health concerns and leading causes of chronic liver disease. In 2022, the World Health Organization estimated that 253 million people were living with chronic hepatitis B infection globally, with 63% of cases occurring in the African and Western Pacific regions^1^. Chronic viral hepatitis led to approximately 1.1 million deaths in 2022, primarily due to cirrhosis and liver cancer^1^. The burden of disease is particularly high in low- and middle-income countries.

A metadata analysis done 2024 by Yirsaw et al.^2^ on pregnant women in east Africa showed a prevalence of 6% of HBV infection. A study conducted in 2024 in Rwanda on patients with non-communicable diseases showed that the prevalence of hepatitis B was 2.1%, while the prevalence of hepatitis C was 6.7% ^3^. Chronic HBV and HCV infections often remain asymptomatic for decades, but can progressively damage the liver, eventually resulting in cirrhosis, liver failure, or hepatocellular carcinoma if left untreated^4–6^

Ultrasound serves as a non-invasive, cost-effective, and widely available tool for assessing liver morphology, detecting complications, and screening for hepatocellular carcinoma (HCC) ^7–11^. Common sonographic findings include hepatomegaly, liver steatosis, liver cirrhosis, and hepatic nodules^4,12–15^. The progression and severity of these findings can vary based on factors such as the duration of infection, coexisting conditions and patient lifestyle^9^.

Sonographic patterns in HBV and HCV infections reflect their distinct pathogenic mechanisms, with HCV demonstrating stronger associations with metabolic manifestations. While both viruses culminate in similar end-stage cirrhotic features (surface nodularity, blunted edges), HCV patients exhibit significantly higher rates of steatosis (79% prevalence) and hepatomegaly (42.9% vs. 32% in HBV), underscoring its pronounced metabolic disruption^13,15^. This metabolic propensity is further evidenced by fatty liver occurring in 11.1% of HBV/HCV co-infections compared to 0% in HBV/hepatitis D virus (HDV) cases^16^.

Older age and male sex have been identified as significant epidemiologic factors impacting the progression and outcomes of viral hepatitis^17^. Studies have found that increasing age is associated with a higher risk of developing cirrhosis and hepatocellular carcinoma in patients with chronic viral hepatitis, likely due to longer duration of infection over time ^18^. Additionally, male gender has consistently been recognized as an independent risk factor for more advanced liver disease and liver-related complications in viral hepatitis patients ^19^. Understanding these patterns in the Rwandan context is essential for developing targeted interventions and improving patient outcomes.

Rwanda has made significant strides in addressing infectious diseases, including hepatitis, through national health policies and programs. The Rwandan Ministry of Health, in collaboration with international partners, has implemented hepatitis screening, vaccination, and management programs of positive diagnosed patients where regular screening of liver damage is among the management^20,21^. Despite these efforts, literature indicates that early detection of liver abnormalities through sonography can significantly improve patient outcomes, highlighting the need for integrating regular sonographic screening into Rwanda’s HBV management protocols. However, the lack of localized data on sonographic patterns and their clinical implications hinders the development of effective guidelines tailored to the Rwandan population.

Several knowledge gaps limit the enhancement of current hepatitis management strategies. These include limited data on types of sonographic findings in HBV and HCV patients in Rwanda and insufficient understanding of the clinical and demographic factors associated with specific sonographic patterns.

This study aims to address these knowledge gaps by identifying sonographic patterns and findings in HBV and HCV patients at Rwanda Military Referral and Teaching Hospital, investigating the influence of social demographic factors (age and gender) on hepatic sonographic findings, and comparing findings in HBV patient with those with HCV.

By providing detailed insights into the sonographic evaluation of HBV and HCV patients, this study will inform clinical practice, enhance diagnostic accuracy, and contribute to the development of tailored management guidelines. Ultimately, it aims to improve patient outcomes and support the integration of sonography into the comprehensive care of hepatitis B and C in Rwanda.

## 2. Methods

### Study design and setting

this study employed a cross-sectional design, involving the collection, analysis, and reporting of primary quantitative data. The research was conducted at the Rwanda Military Referral and Teaching Hospital (RMRTH), a national referral and teaching hospital located in Kigali City, Rwanda. RMRTH receives hepatitis B and C patients from across Rwanda, largely due to its connection with the Ministry of Defense’s outreach screening program^22^.

The radiology department at RMRTH conducts an average of 1,920 liver ultrasounds annually for hepatitis B and C patients, following the hospital’s mandatory ultrasound protocol for hepatitis management. Equipped with advanced imaging technology and staffed by five radiologists and six sonographers, the department provides a high-volume, standardized setting ideal for studying sonographic liver patterns.

### Study Population and sampling

All patients with a confirmed diagnosis of hepatitis B or hepatitis C infection who underwent ultrasound scans were the source population. The study excluded participants with known chronic liver diseases attributable to causes other than HBV/HCV infection. The sample size was determined using a modified Cochran’s formula for finite population^23^. With an average population of 640 within 4 months. We considered a 95% confidence level with a margin of error equal to 0.05. This yielded a required sample size of 240 participants.

A consecutive sampling approach was used. Averagely five patients per day diagnosed with chronic hepatitis B or C who presented to the ultrasound department during the study period were invited to participate in this study.

### Data collection

Patient age and gender as demographic data were obtained. B-mode ultrasound findings of the liver were recorded using SIEMENS ACUSON Sequoia with a combination of an ultrasound machine with a 1.0 to 5.7 MHz curved array transducer and a 2.9 to 9.9 MHz linear array transducer.

The following sonographic data were collected and filled in the data collection sheet:

- Liver size by measuring the right liver lobe at the MCL: the size between 13 and 16cm was considered as normal, the liver size below 13cm was considered shrunken and the size above 16cm was considered as hepatomegaly
- Liver echogenicity: assessed by comparing the liver and renal cortex. The echogenicity was considered normal when the liver is isoechoic or slightly hyperechoic to the renal cortex. Increased echogenicity was noted when the liver is relatively hyperechoic to the renal cortex. Reduced echogenicity was noted when the liver echogenicity was hypoechoic to the renal cortex.
- Liver morphological change suggestive of liver fibrosis was assessed using the Ultrasonographic Scoring System (USSS) described by Nishiura et al.^11^, which semi-quantitatively grades sonographic features suggestive of fibrosis, based on three parameters: liver edge (0=sharp, 1=mildly blunted, 2=blunted), liver surface (0=smooth, 1=mildly irregular, 2=irregular, 3=highly irregular), and liver parenchymal texture (0=fine,1=mildly coarse, 2=coarse, 3=highly coarse). Each parameter was scored separately for the right and left lobes, and the average score was used to obtain the final value. A score of 0 indicated no abnormality on high frequency probe, score 1 indicated mild abnormality detected only by high frequency probe, score 2 indicated moderate abnormality detected by low frequency probe, and score 3 indicated severe abnormality clearly confirmed by low frequency probe (**Figure 3 and Figure 4**). Scores of 0–3 were categorized as no or mild morphological change, 3.5–4.5 as moderate change, 5–6 as advanced change, and 6.5–8 as marked morphological change consistent with cirrhotic morphology.
- Presence of a liver nodule: The Li-RADS classification was used to categorize detected nodules, with Li-RADS 1 indicating benign nodules and Li-RADS 2 and 3 indicating suspicious liver nodules^24^.

### Statistical analysis

Data analysis was conducted using SPSS version 26.0. Descriptive statistics were used to summarize the demographic characteristics of participants and the prevalence of sonographic findings.

To identify prevalent sonographic patterns and findings, frequencies and percentages were calculated for each sonographic pattern and finding. To explore associations between sonographic findings and clinical/demographic factors, chi-square tests were used for categorical variables and independent t-tests for continuous variables. A p-value < 0.05 was considered statistically significant. Results were presented using tables and graphs as appropriate.

### Ethical considerations

Ethical approval for this study was obtained from the Institutional Review Board of the Rwanda Military Teaching Hospital (Approval No. 552/RMRTH/COMDT/2024) on 9 December 2024, prior to the commencement of data collection. Written informed consent was obtained from all participants after explaining the study purpose, procedures, and their right to withdraw at any time without consequence. All collected data were handled confidentially, with participants assigned unique identification codes to ensure anonymity. Access to the dataset was restricted to the research team only, and no identifying personal information was disclosed in the final report or publication..

## 3. Results

### 3.1. Social demographic characteristics

Findings presented in this section are out of 240 ultrasound examinations performed on 120 participants with HBV and 120 participants with HCV. Over than a half of participants: 137 (57.1%) were male, while 103 (42.9%) were female.

The average age of the participants was 52.7 ± 17.9 years, with an observed age range of 20 to 93years (**Table 1**).

**Table 1.**
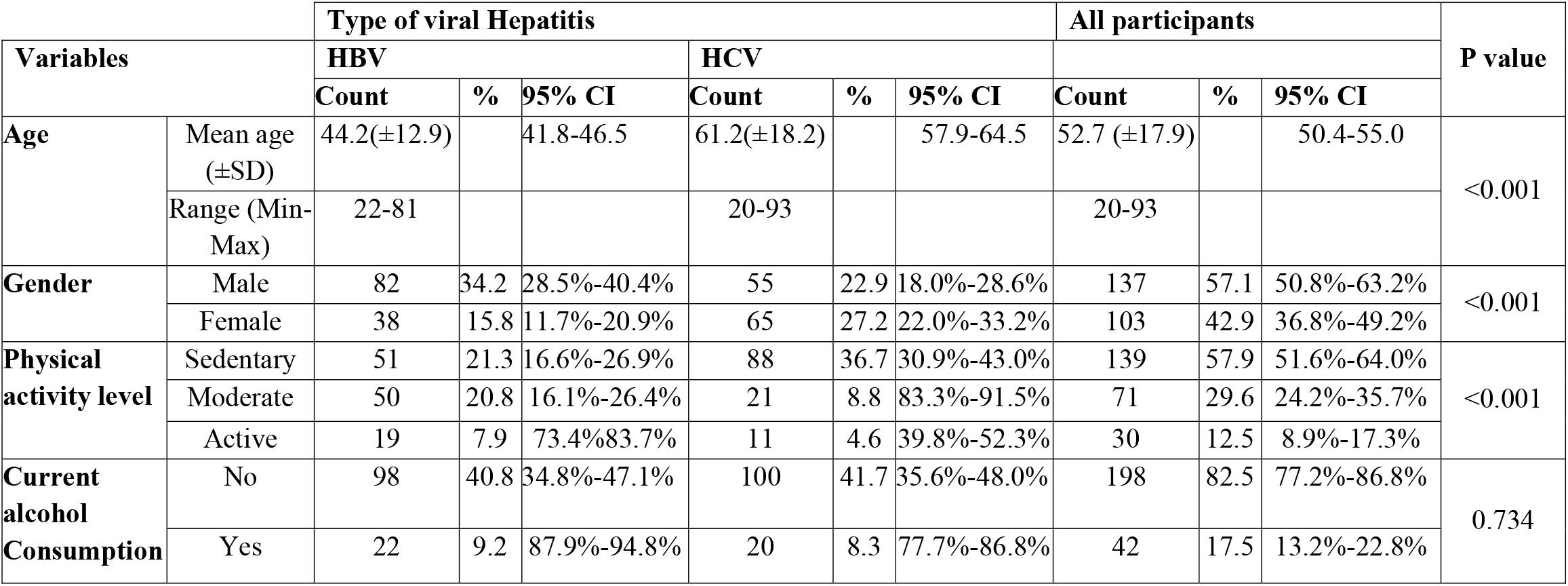
Social demographic characteristics of participants.

|  |  | Type of viral Hepatitis |  |  |  |  |  | All participants |  |  | P value |
| --- | --- | --- | --- | --- | --- | --- | --- | --- | --- | --- | --- |
| Variables |  | HBV |  |  | HCV |  |  |  |  |  |  |
|  |  | Count | % | 95% CI | Count | % | 95% CI | Count | % | 95% CI |  |
| Age | Mean age (±SD) | 44.2(±12.9) |  | 41.8-46.5 | 61.2(±18.2) |  | 57.9-64.5 | 52.7 (±17.9) |  | 50.4-55.0 | <0.001 |
|  | Range (Min-Max) | 22-81 |  |  | 20-93 |  |  | 20-93 |  |  |  |
| Gender | Male | 82 | 34.2 | 28.5%-40.4% | 55 | 22.9 | 18.0%-28.6% | 137 | 57.1 | 50.8%-63.2% | <0.001 |
|  | Female | 38 | 15.8 | 11.7%-20.9% | 65 | 27.2 | 22.0%-33.2% | 103 | 42.9 | 36.8%-49.2% |  |
| Physical activity level | Sedentary | 51 | 21.3 | 16.6%-26.9% | 88 | 36.7 | 30.9%-43.0% | 139 | 57.9 | 51.6%-64.0% | <0.001 |
|  | Moderate | 50 | 20.8 | 16.1%-26.4% | 21 | 8.8 | 83.3%-91.5% | 71 | 29.6 | 24.2%-35.7% |  |
|  | Active | 19 | 7.9 | 73.4%83.7% | 11 | 4.6 | 39.8%-52.3% | 30 | 12.5 | 8.9%-17.3% |  |
| Current alcohol Consumption | No | 98 | 40.8 | 34.8%-47.1% | 100 | 41.7 | 35.6%-48.0% | 198 | 82.5 | 77.2%-86.8% | 0.734 |
|  | Yes | 22 | 9.2 | 87.9%-94.8% | 20 | 8.3 | 77.7%-86.8% | 42 | 17.5 | 13.2%-22.8% |  |

Regarding physical activity levels, the majority of participants were sedentary, accounting for 139 (57.9%) of the total study population. Among HBV participants, 51 (21.3%) were sedentary, 50 (20.8%) had moderate physical activity, and 19 (7.9%) were active. In contrast, HCV participants showed a different distribution, with 88 (36.7%) being sedentary, 21 (8.8%) having moderate physical activity, and only 11 (4.6%) being active.

Alcohol consumption was relatively low among the participants, with 42 (17.5%) reporting current alcohol use. Specifically, 22 (9.2%) of HBV participants and 20 (8.3%) of HCV participants reported current alcohol consumption. The majority of participants (198, 82.5%) reported that they were not currently consuming alcohol.

Most social demographic characteristics showed statistically significant differences between HBV and HCV groups, with p-values less than 0.001 for age, gender, and physical activity level, except for alcohol consumption (p=0.734), which did not show a significant difference between the two hepatitis types. Detailed social demographic characteristics are highlighted in **Table 1**

### 3.2. Hepatic sonographic patterns among participants

While the majority of participants showed normal sonographic liver patterns (**Table 2, Figure1)**. Among the abnormal sonographic patterns, the most frequent findings were coarse echotexture, surface nodularity, and blunt liver edge, features commonly suggestive of early fibrotic change. Using the Ultrasonographic Scoring, which combines the degree of echotexture coarseness, surface nodularity, and edge blunting. Most participants (78.3%) fell within the no or mild morphological change range, while about one-fifth (21.7%) showed moderate to marked morphological change, including 10% with features consistent with cirrhosis (**Figure2**). A smaller proportion of participants showed altered right lobe size and increased echogenicity, while sonographic features consistent with liver nodules (LI-RADS 2–3) were less frequent.

**Table 2.** Distribution of ultrasound liver patterns among study participants.

| Ultrasound patterns |  | Count | % | 95% CI |
| --- | --- | --- | --- | --- |
| Liver echogenicity | Normal | 219 | 91.3% | 87.1-94.2% |
|  | Increased | 21 | 8.8% | 5.8%-13.1% |
| Liver echotexture | Homogeneous | 182 | 75.8% | 70.0%-80.2% |
|  | Coarse | 58 | 24.2% | 19.2%-30.0% |
| Liver surface | Smooth | 183 | 76.3% | 70.5%-81.2% |
|  | Nodular | 57 | 23.8% | 18.8%-29.5% |
| Liver edge | Sharp | 188 | 78.3% | 72.7%-83.0% |
|  | Blunt | 52 | 21.7% | 17.0%-28.3% |
| Right lobe size | Normal size | 205 | 85.4% | 80.4%-89.4% |
|  | Reduced size | 28 | 11.7% | 8.2%-16.4% |
|  | Increased size | 7 | 2.9% | 1.4%-5.9% |
| Liver Nodule | No nodule | 222 | 92.5% | 88.5%-95.2% |
|  | Li-RADS 1 nodule | 6 | 2.5% | 1.2%-5.3% |
|  | Li-RADS 2 or 3 nodule | 12 | 5.0% | 2.9%-8.5% |

**Figure 1:**
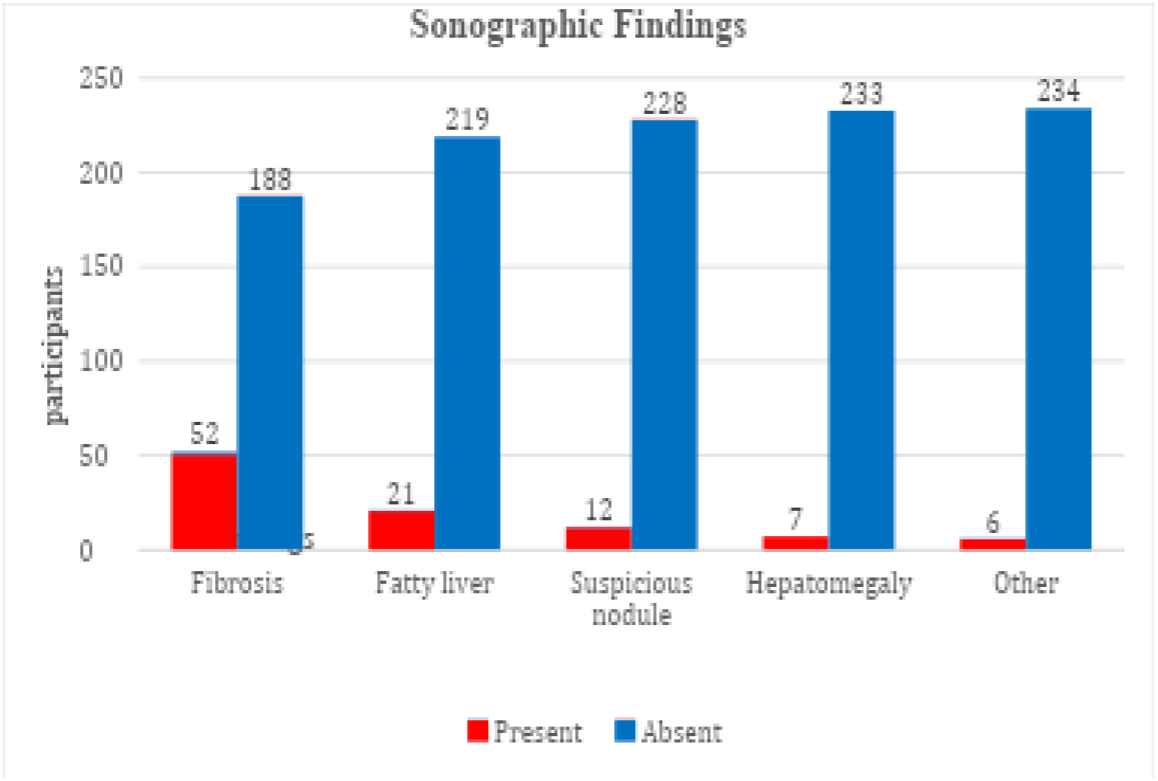
Distribution of sonographic findings among participants.

**Figure 2:**
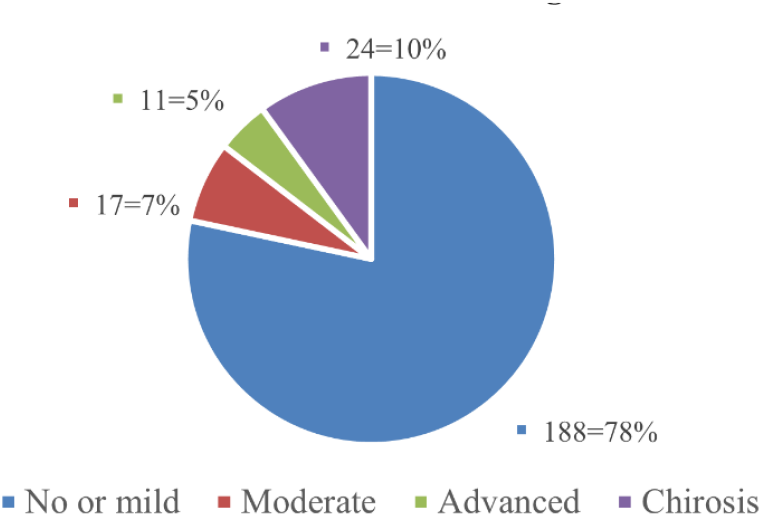
Distribution of sonographic morphological change patterns suggestive of fibrosis among study participants.

**Figure 3:**
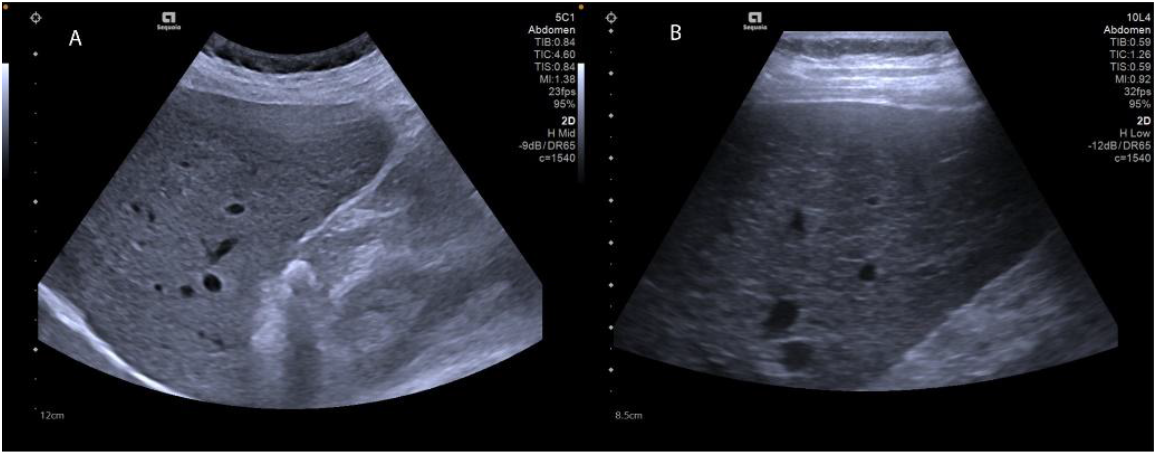
Ultrasound images of a liver from a female participant in her 50’s with HCV. (A) Curved array low-frequency probe image demonstrating coarse echotexture and blunted edge of the right lobe. (B) Linear array probe image showing coarse echotexture and mildly irregular surface. These findings were categorized as advanced morphological change.

**Figure 4:**
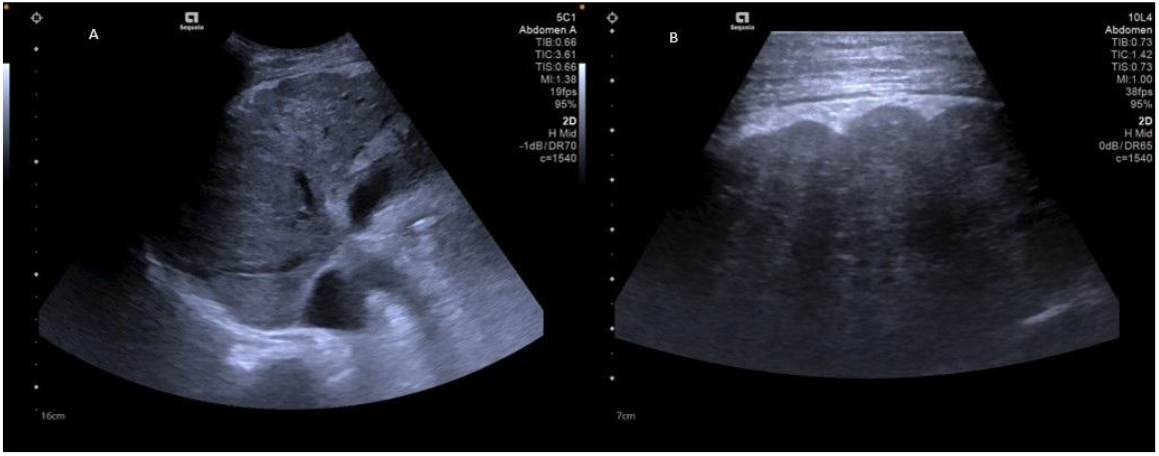
Ultrasound images of a liver from a male participant in his 50’s with HCV. (A) Curved array low-frequency probe image demonstrating highly coarse echotexture. (B) Linear array probe image showing highly coarse echotexture and highly irregular surface. These findings were categorized as marked morphological change consistent with cirrhotic appearance.

When findings were categorized by diagnostic pattern, 21.7% of participants showed sonographic patterns suggestive of fibrosis, 8.8% showed features of fatty liver, 5.0% had suspicious liver nodules, 2.9% hepatomegaly, and 2.5% other hepatic abnormalities.

### 3.3. Comparison of hepatic sonographic findings in HBV and HCV patients

Fibrosis patterns was the only statistically significant finding between the two hepatitis types (p = 0.012), occurring in 7.5% of HBV and 14.2% of HCV patients. sonographic patterns of fatty liver, hepatomegaly, and other liver findings showed no significant variation (p > 0.05). Suspicious nodules were more frequent in HCV (3.8%) than HBV (1.3%) cases but did not reach statistical significance (p = 0.076) (**Table 3**).

**Table 3.** Crosstabulation comparing type of hepatitis and liver findings.

| Findings |  | HBV |  | HCV |  | Total |  | P value |
| --- | --- | --- | --- | --- | --- | --- | --- | --- |
|  |  | Count | % | Count | % | Count | % |  |
| Fibrotic patterns | Yes | 18 | 7.5 | 34 | 14.2 | 52 | 21.7 | 0.012 |
|  | No | 102 | 42.5 | 86 | 35.8 | 188 | 78.3 |  |
|  | Total | 120 | 50 | 120 | 50 | 240 | 100 |  |
| Fatty liver | Yes | 10 | 4.2 | 11 | 4.6 | 21 | 8.8 | 0.819 |
|  | No | 110 | 45.8 | 109 | 45.4 | 219 | 91.3 |  |
|  | Total | 120 | 50 | 120 | 50 | 240 | 100 |  |
| Suspicious liver nodule | Yes | 3 | 1.3 | 9 | 3.8 | 12 | 5 | 0.076 |
|  | No | 117 | 48.8 | 111 | 46.3 | 228 | 95 |  |
|  | Total | 120 | 50 | 120 | 50 | 240 | 100 |  |
| Hepatomegaly | Yes | 2 | 0.8 | 5 | 2.1 | 7 | 2.9 | 0.250 |
|  | No | 118 | 49.2 | 115 | 47.9 | 233 | 97.1 |  |
|  | Total | 120 | 50 | 120 | 50 | 240 | 100 |  |
| Other liver findings | Yes | 4 | 1.7 | 2 | 0.8 | 6 | 2.5 | 0.408 |
|  | No | 116 | 48.3 | 118 | 49.2 | 234 | 97.5 |  |
|  | Total | 120 | 50 | 120 | 50 | 240 | 100 |  |

### 3.4. Association between sociodemographic factors and hepatic sonographic findings

As shown in Table 4, Age groups demonstrated a strong correlation with fibrotic patterns, with a highly significant p-value of <0.001. The prevalence of morphological change increased markedly with age: no cases in the ≤30 age group, rising to 5.4% in the ≥76 age group. The 61-75 age group showed the highest percentage at 10% of sonographic features suggesting liver fibrosis.

**Table 4.** Cross tabulation between age groups and liver ultrasound findings.

| Ultrasound findings |  | Age group |  |  |  |  |  |  |  |  |  |  |  | P value |
| --- | --- | --- | --- | --- | --- | --- | --- | --- | --- | --- | --- | --- | --- | --- |
|  |  | ≤ 30 |  | 31-45 |  | 46-60 |  | 61-75 |  | ≥ 76 |  | Total |  |  |
|  |  | Count | % | Count | % | Count | % | Count | % | Count | % | Count | % |  |
| Fibrotic patterns | Yes | 0 | 0 | 3 | 1.3 | 12 | 5 | 24 | 10 | 13 | 5.4 | 52 | 22 | <0.001 |
|  | No | 28 | 12 | 69 | 29 | 47 | 20 | 22 | 9.2 | 22 | 9.2 | 188 | 78 |  |
|  | Total | 28 | 12 | 72 | 30 | 59 | 25 | 46 | 19 | 35 | 15 | 240 | 100 |  |
| Fatty liver | Yes | 0 | 0 | 5 | 2.1 | 10 | 4.2 | 5 | 2.1 | 1 | 0.4 | 21 | 8.8 | 0.457 |
|  | No | 28 | 12 | 67 | 28 | 49 | 20 | 41 | 17 | 34 | 14 | 219 | 91 |  |
|  | Total | 28 | 12 | 72 | 30 | 59 | 25 | 46 | 19 | 35 | 15 | 240 | 100 |  |
| Suspicious liver nodule | Yes | 0 | 0 | 3 | 1.3 | 5 | 2.1 | 3 | 1.3 | 1 | 0.4 | 12 | 5 | 0.457 |
|  | No | 28 | 12 | 69 | 29 | 54 | 23 | 43 | 18 | 34 | 14 | 228 | 95 |  |
|  | Total | 28 | 12 | 72 | 30 | 59 | 25 | 46 | 19 | 35 | 15 | 240 | 100 |  |
| Hepatomegaly | Yes | 2 | 0.8 | 1 | 0.4 | 1 | 0.4 | 2 | 0.8 | 1 | 0.4 | 7 | 2.9 | 0.557 |
|  | No | 26 | 11 | 71 | 30 | 58 | 24 | 44 | 18 | 34 | 14 | 233 | 97 |  |
|  | Total | 28 | 12 | 72 | 30 | 59 | 25 | 46 | 19 | 35 | 15 | 240 | 100 |  |
| Other liver findings | Yes | 0 | 0 | 0 | 0 | 4 | 1.7 | 1 | 0.4 | 1 | 0.4 | 6 | 2.5 | 0.134 |
|  | No | 28 | 12 | 72 | 30 | 55 | 23 | 45 | 19 | 34 | 14 | 234 | 98 |  |
|  | Total | 28 | 12 | 72 | 30 | 59 | 25 | 46 | 19 | 35 | 15 | 240 | 100 |  |

Fatty liver and suspicious liver nodules showed minimal variation across age groups, with non-significant p-values of 0.457. Hepatomegaly and other liver findings also did not demonstrate statistically significant correlations with age groups, with p-values of 0.557 and 0.134, respectively.

Fibrotic patterns showed a statistically significant difference between males and females (p=0.006). While 8.8% of males had patterns of fibrosis, 12.9% of females were affected. (**Table 5**)

**Table 5.** Crosstabulation between Gender and liver findings.

| Findings |  | Gender |  |  |  |  |  | P value |
| --- | --- | --- | --- | --- | --- | --- | --- | --- |
|  |  | Male |  | Female |  | Total |  |  |
|  |  | Count | % | Count | % | Count | % |  |
| Fibrotic patterns | Yes | 21 | 8.8 | 31 | 12.9 | 52 | 21.7 | 0.006 |
|  | No | 116 | 48.3 | 72 | 30 | 188 | 78.3 |  |
|  | Total | 137 | 57.1 | 103 | 42.9 | 240 | 100 |  |
| Fatty liver | Yes | 13 | 5.4 | 8 | 3.3 | 21 | 8.8 | 0.640 |
|  | No | 124 | 51.7 | 95 | 39.6 | 219 | 91.3 |  |
|  | Total | 137 | 57.1 | 103 | 42.9 | 240 | 100 |  |
| Suspicious liver nodule | Yes | 7 | 2.9 | 5 | 2.1 | 12 | 5 | 0.928 |
|  | No | 130 | 54.2 | 98 | 40.8 | 228 | 95 |  |
|  | Total | 137 | 57.1 | 103 | 42.9 | 240 | 100 |  |
| Hepatomegaly | Yes | 6 | 2.5 | 1 | 0.4 | 7 | 2.9 | 0.120 |
|  | No | 131 | 54.6 | 102 | 42.5 | 233 | 97.1 |  |
|  | Total | 137 | 57.1 | 103 | 42.9 | 240 | 100 |  |
| Other liver findings | Yes | 1 | 0.4 | 5 | 2.1 | 6 | 2.5 | 0.043 |
|  | No | 136 | 56.7 | 98 | 40.8 | 234 | 97.5 |  |
|  | Total | 137 | 57.1 | 103 | 42.9 | 240 | 100 |  |

Other liver findings also showed a statistically significant gender difference (p=0.043), with females (2.1%) more frequently presenting such findings compared to males (0.4%). Hepatomegaly approached but did not reach statistical significance (p=0.120), with females showing lower occurrence.

Fatty liver and suspicious liver nodules demonstrated no statistically significant gender differences, with p-values of 0.640 and 0.928, respectively.

## 4. Discussion

### 4.1. Sonographic patterns and findings in HBV and HCV patients

The most frequent abnormal ultrasound findings were coarse echotexture, surface nodularity, and blunt liver edge, features commonly associated with chronic parenchymal change. These patterns align with established ultrasound markers of chronic liver disease described by Colli et al.^14^, who identified liver surface nodularity as a reliable sign of advanced fibrosis. The relatively high frequency of these changes suggests that many patients in our cohort had already developed significant parenchymal alteration by the time of this study.

Patterns suggestive of fibrosis was seen in 21.7% of participants, lower than the 39% reported by Ali et al.^13^. Variations in population characteristics, disease duration, and access to antiviral therapy may account for these differences.

Fatty liver was noted in 8.8% of participants, a much lower rate than the 79% reported by Wang et al.^15^ in a Taiwanese cohort, possibly reflecting differing metabolic risk factors and lifestyle profiles.

Hepatomegaly was relatively uncommon (2.9%), while right-lobe volume reduction was observed in 11.7%, suggesting that several patients were at advanced stages, where fibrosis results in shrinkage rather than enlargement.

Suspicious liver nodules were found in 5% of participants, underscoring the importance of regular ultrasound surveillance for early detection of hepatocellular carcinoma, consistent with global findings^25–27^

### 4.2. Comparison of hepatic sonographic findings in HBV Vs. HCV patients

Regarding the comparison of the hepatic sonographic findings in patients with HBV and those with HCV. The most observed difference was the significantly higher prevalence of liver fibrosis patterns in HCV participants compared to HBV participants (28.3% vs. 15.0%, (χ^2^ = 6.285, df = 1, p=0.012, Cramer’s V= 0162). This finding aligns with the natural history of these infections, as HCV more commonly leads to chronic infection than HBV, with approximately 80% of HCV-infected individuals developing chronic hepatitis compared to only 5-10% of adults infected with HBV^12^.

The higher prevalence of fibrosis patterns in HCV patients may also be related to the older age of this group, as noted previously. With a mean age of 61.18 years for HCV patients versus 44.15 years for HBV patients, the HCV group likely had longer disease duration, allowing more time for fibrosis development. This interaction between virus type and age underscores the complex interplay of factors influencing liver disease progression.

No statistically significant differences were observed between HBV and HCV patients regarding fatty liver, suspicious liver nodules, hepatomegaly, or other liver findings. This suggests that while the progression of liver morphological change may differ between these viral infections, the spectrum of other hepatic manifestations remains relatively similar. This is consistent with the shared pathways in causing liver damage described by Petruzziello^26^, despite HBV and HCV being different types of viruses.

### 4.3. Correlation between sociodemographic factors and hepatic sonographic findings

Our analysis revealed a statistically significant association between age and patterns of liver fibrosis (χ^2^ = 50.962, df = 4, p < 0.001), with a moderate to strong effect size (Cramer’s V = 0.461). The prevalence of fibrotic patterns increased steadily with age, from 0% in those ≤30 years to 52.2% in those aged 61-75 years, before slightly decreasing to 37.1% in participants ≥76 years. This age-related pattern is consistent with the natural history of viral hepatitis, where increasing age typically correlates with longer duration of infection and greater cumulative liver damage. Nguyen et al.^18^ similarly found that increasing age is associated with higher risk of developing cirrhosis and hepatocellular carcinoma in chronic viral hepatitis patients.

With respect to gender, female participants demonstrated a significantly higher prevalence of fibrotic patterns compared to males (30.1% vs. 15.3%, p=0.006). This finding contrasts with Yang et al^19^, who identified male gender as an independent risk factor for more advanced liver disease in viral hepatitis patients. This unexpected result may reflect confounding factors not accounted for in our analysis, such as differing duration of infection, or variations in healthcare-seeking behavior between genders in our Rwandan population.

The demographic characteristics of our study population showed notable differences, with males predominating in the HBV group (68.3%) and females slightly predominating in the HCV group (54.2%). This pattern aligns with epidemiological data showing HBV infections are often more common in males^17^. Additionally, HCV patients were substantially older (mean age 61.18 years) than HBV patients (mean age 44.15 years), which may reflect different transmission patterns and epidemic histories of these viruses in Rwanda.

This study has a few limitations such as the absence of histological or elastographic correlation limits the ability to confirm the exact fibrosis stage. Additionally, possible confounders such as treatment history and comorbidities were not controlled. These factors should be considered when interpreting the findings.

The significant association between older age, female gender, and morphological liver changes highlights the importance of targeted monitoring strategies. The gender difference observed may suggest variations in infection duration or healthcare-seeking behavior, warranting further local investigation.

Given its accessibility, ultrasound remains a practical first-line tool for identifying and following up parenchymal liver changes in resource-limited settings. Future studies integrating ultrasound with elastography or biochemical markers could improve the accuracy and validation of ultrasound-based liver assessment in African clinical contexts.

## 5. Conclusion

Ultrasound revealed distinct morphological liver changes among patients with hepatitis B and C, most commonly coarse echotexture, surface nodularity, and blunted edge—features suggestive of parenchymal alteration. These sonographic features were more frequent in HCV and in older participants, reflecting typical patterns of chronic liver disease progression.

Interestingly, female participants showed a higher prevalence of parenchymal morphological changes, a finding that may reflect context-specific factors such as differences in infection duration or healthcare access and warrants further investigation.

Although ultrasound alone cannot determine histological fibrosis stage, it remains a valuable and practical tool for identifying and monitoring morphological liver changes in resource-limited settings.

Future studies in Rwanda and similar contexts could combine ultrasound with elastography or biochemical assessment to validate and expand on these findings, including possible gender-related variations.

## Data Availability

All data generated and analyzed during this study are available from the corresponding author upon reasonable request.

## References

1. World Health Organization. Global Hepatitis Report 2024 Action for Access in Low- and Middle-Income Countries. World Health Organization; 2024.

2. Yirsaw BG, Agimas MC, Alemu GG, Tesfie TK, Derseh NM, Abuhay HW, et al. Prevalence of Hepatitis B virus infection and its determinants among pregnant women in East Africa: Systematic review and Meta-analysis. PLoS One. 2024;19.

3. Musafiri T, Kamali I, Kayihura C, de la Paix Gakuru J, Nyirahabihirwe F, Nizeyimana E, et al. Prevalence of hepatitis B and C infection and linkage to care among patients with Non-Communicable Diseases in three rural Rwandan districts: a retrospective cross-sectional study. BMC Infect Dis. 2024;24.

4. Lampertico P, Agarwal K, Berg T, Buti M, Janssen HLA, Papatheodoridis G, et al. EASL 2017 Clinical Practice Guidelines on the management of hepatitis B virus infection. J Hepatol. 2017;67:370–98.

5. Seeger C, Mason WS. Molecular biology of hepatitis B virus infection. Vols. 479–480, Virology. Academic Press Inc.; 2015. p. 672–86.

6. Scheel TKH, Rice CM. Understanding the hepatitis C virus life cycle paves the way for highly effective therapies. Vol. 19, Nature Medicine. 2013. p. 837–49.

7. Ferraioli G. Review of liver elastography guidelines. Journal of Ultrasound in Medicine. 2019;38:9–14.

8. Choong CC, Venkatesh SK, Siew EPY. Accuracy of Routine Clinical Ultrasound for Staging of Liver Fibrosis. J Clin Imaging Sci. 2012;2:58.

9. Bohte AE, De Niet A, Jansen L, Bipat S, Nederveen AJ, Verheij J, et al. Non-invasive evaluation of liver fibrosis: A comparison of ultrasound-based transient elastography and MR elastography in patients with viral hepatitis B and C. Eur Radiol. 2014;24:638–48.

10. Gerstenmaier JF, Gibson RN. Ultrasound in chronic liver disease. Insights Imaging [Internet]. 2014;5:441–55. Available from: http://link.springer.com/10.1007/s13244-014-0336-2

11. Nishiura T, Watanabe H, Ito M, Matsuoka Y, Yano K, Daikoku M, et al. Ultrasound evaluation of the fibrosis stage in chronic liver disease by the simultaneous use of low and high frequency probes. British Journal of Radiology. 2005;78:189–97.

12. Westbrook RH, Dusheiko G. Natural history of hepatitis C. Vol. 61, Journal of Hepatology. Elsevier B.V.; 2014. p. S58–68.

13. Ali ARN, Qayyum S, Farooq SMY, Jillani A, Nauman M, Ghazi AR, et al. Sonographic Findings in patients of Hepatitis B & C. Pakistan Journal of Medical and Health Sciences. 2022;16:11–4.

14. Colli A, Fraquelli M, Andreoletti M, Marino B, Zuccoli E, Conte D. Severe liver fibrosis or cirrhosis: Accuracy of US for detection - Analysis of 300 cases. Radiology. 2003;227:89–94.

15. Wang TJ, Chen MY, Lin YC, Chiu WN, Huang TJ, Weng HH. High prevalence of fatty liver and its association with metabolic syndrome among rural adults with chronic hepatitis C: Implications for primary healthcare. BMC Public Health. 2024;24.

16. Ebrahimzadeh A, Alemzadeh E, Mohammadifard M, Askari P. Comparison of serologic and sonographic findings in patients with HBV/HDV and HBV/HCV Infections. Vol. 1, Research in Health & Medical Sciences Original Article. 2023.

17. El-Serag HB. Epidemiology of viral hepatitis and hepatocellular carcinoma. Gastroenterology. 2012;142.

18. Nguyen MH, Wong G, Gane E, Kao JH, Dusheiko G. Hepatitis B Virus: Advances in Prevention, Diagnosis, and Therapy [Internet]. 2020. Available from: http://cmr.asm.org/

19. Yang JD, Kim WR, Coelho R, Mettler TA, Benson JT, Sanderson SO, et al. Cirrhosis Is Present in Most Patients With Hepatitis B and Hepatocellular Carcinoma. Clinical Gastroenterology and Hepatology. 2011;9:64–70.

20. Mbituyumuremyi A, Van Nuil JI, Umuhire J, Mugabo J, Mwumvaneza M, Makuza JD, et al. Controlling hepatitis c in rwanda: A framework for a national response. Bull World Health Organ. 2018;96:51–8.

21. Makuza JD, Tuyishime A, Janjua N, Gupta N. HBV elimination in sub-Saharan Africa: Rwanda’s approach to health system integration. Lancet Gastroenterol Hepatol [Internet]. 2022;7:511–2. Available from: 10.1016/S2468-1253(22)00134-0

22. King K, Paul BJ, Boniface N, Pacifique K, Janviere M, Fidele B, et al. Medical citizen outreach programs as Rwanda Defense Force homegrown solution for health challenges in Rwanda [Internet]. Vol. 1, Public Health Bul. 2019. Available from: https://www.rwandapublichealthbulletin.org

23. Cochran WG. Sampling techniques. Wiley; 1977.

24. Kamaya A, Fetzer DT, Seow JH, Burrowes DP, Choi HH, Dawkins AA, et al. LI-RADS US Surveillance Version 2024 for Surveillance of Hepatocellular Carcinoma: An Update to the American College of Radiology US LI-RADS. Vol. 313, Radiology. 2024. p. e240169.

25. Ringehan M, McKeating JA, Protzer U. Viral hepatitis and liver cancer. Vol. 372, Philosophical Transactions of the Royal Society B: Biological Sciences. Royal Society Publishing; 2017.

26. Petruzziello A. Epidemiology of Hepatitis B Virus (HBV) and Hepatitis C Virus (HCV) Related Hepatocellular Carcinoma. Open Virol J. 2018;12:26–32.

27. Heller MT, Tublin ME. The role of ultrasonography in the evaluation of diffuse liver disease. Vol. 52, Radiologic Clinics of North America. W.B. Saunders; 2014. p. 1163–75.

